# Comprehensive Histopathology Imaging in Pancreatic Biopsies: High Definition Infrared Imaging with Machine Learning Approach

**DOI:** 10.1101/2022.07.01.22277130

**Authors:** Danuta Liberda, Paulina Koziol, Tomasz P. Wrobel

**Affiliations:** Solaris National Synchrotron Radiation Centre, Jagiellonian University, Czerwone Maki 98, 30-392 Krakow, Poland; Institute of Physics, Jagiellonian University, Lojasiewicza 11, 30-348 Krakow, Poland

**Keywords:** Pancreatic cancer, infrared imaging, machine learning, high definition, quantum cascade laser

## Abstract

Infrared (IR) based histopathology offers a new paradigm in looking at tissues and can provide a complimentary information source for more classical histopathology, which makes it a noteworthy tool given possible clinical application. The goal of this study is to build a robust machine learning model using IR imaging of pancreatic cancer histopathology on a single pixel level. In this article, we report a pancreatic cancer classification model based on data from over 600 biopsies (coming from 250 patients) imaged with IR diffraction-limited spatial resolution. To fully research model’s classification ability, we measured tissues in Standard and High Definition using two optical setups. This forms one of the largest IR datasets analyzed up to now, with almost 700 million spectra of different tissue types. The first classification model, based on six tissue classes, created for comprehensive histopathology achieved AUC values on the pixel (tissue) level above 0.95. We successfully developed a comprehensive histopathology digital staining model for pancreatic tissues based on biochemical information extracted from IR spectra.

## Introduction

Pancreatic ductal adenocarcinoma (PDAC), the most common pancreatic cancer type, accounts for less than 3% of all cancers, but it remains the third leading cause of cancer-related deaths, both for men and women in the United States with 53,000 estimated new cases and an 80% death rate in one year [1]. Furthermore, PDAC is projected to become the second cause of cancer-related death by 2030 [2]. The dreadful prognosis of patients with this disease, including a less than 8% 5-year survival after diagnosis, is due to the lack of early symptoms and/or specific biomarkers for early diagnosis and the paucity of available chemotherapy. In PDAC, three types of biomarkers are desirable: those that help in the detection of the disease onset (diagnosis biomarkers); those that predict responses to treatments (predictive biomarkers) and those that forecast the likely course of the disease, including survival and recurrence patterns (prognosis biomarkers).

Infrared (IR) based histopathology offers a new paradigm in looking at tissues and can provide a complementary information source for more classical histopathology as being label-free and non-destructive [1,2]. Especially, correct understanding of the involved inflammation, fibrosis and neoplasia is required for a method to be successful. IR histopathology is being developed for an increasing number of cancer types, e.g. breast [3], colon [4], lung [5], brain [6] prostate [7], esophagus [8] - highlighting the increasing potential of the method. The technological developments, especially on the speed and spatial resolution front, make this approach feasible in the clinic [9]. The key to the optimization of IR measurements and reducing analysis time is the spectral information reduction: whether it is selection of specific spectroscopic bands [10] or significant reduction of spectral resolution [11]. Both conditions can be met using Quantum Cascade Lasers (QCL) based microscopes. It has been reported that a QCL data collection needed for classification of a centimeter-scale size biopsies could be done in the order of minutes [11,12] with a commercially available Spero-QT (Daylight Solutions, CA, USA) microscope. However, to optimize a method for a fast screening or imaging modality, a proper foundation needs to be laid beforehand even after initial proof of concept studies such as this one [13,14].

Due to the Rayleigh criterion, spatial resolution in IR imaging depends on the wavelength and objective numerical aperture (NA). In this study we applied two optical setups:

- 15x objective with 0.4 NA, giving a projected pixel size of 2.7 μm,
- 36x objective with 0.5 NA, giving a projected pixel size of 1.1 μm.

Considering under-sampling in high wavenumber region (above 2314 cm^-1^) for applied 15x objective, data measured with this optical setup are termed Standard Definition (SD). In the case of the 36x objective, we are not limited in the measured spectral range (3850-900 cm^-1^) by the Rayleigh criterion, therefore, data has High Definition (HD) label. In our previous research [15] we showed that HD data reveal small tissue components like thin fibers or blood cells squeezed in the vessels. However, SD gives more homogenous predictions within tissue classes while also being faster than HD measurements. In this study, we applied both definitions to check their influence on classification on the pixel (tissue) and patient level. It is especially interesting in the case of inflammations that are composed of lymphocytes (size of 7-8 μm).

In this article we report such results for pancreatic cancer histopathology based on FT-IR imaging and machine learning, providing a classification model based on data from over 600 biopsies (coming from 250 patients) imaged with IR diffraction-limited spatial resolution. This forms one of the largest IR datasets analyzed up to now, with almost 700 million spectra of different tissue types.

## Results and discussion

### Patient pathology recognition methodology

This research was performed using Tissue Micro Arrays (TMAs), which are typically applied in studies on tissue composition and differentiation. This is caused by the high variability of TMAs due to many biopsies that are taken from patients differentiated by age, sex, pathology diagnosis, grade, etc.. After the typical procedure of needle biopsies collection, to prevent their damage they are embedded with paraffin and cut into thin 5 μm slices (Figure 1 A). In this research, we used two neighboring slices. Pathology recognition needs to be done based on histopathological annotations performed by an experienced histopathologist (Figure 1B). Therefore, the first TMA slice is H&E (Hematoxylin and Eosin) stained and imaged with a visible light microscope. The second slice is imaged with an IR spectrometer. The result of this measurement is data in the form of a cube containing IR images in the X and Y plane, and spectra in the Z direction (Figure 1C). Based on histopathological annotations mentioned before corresponding regions are marked on the IR image – spectra (after preprocessing) that come from this area are used for final histopathological model creation. This model in the next step is applied to digitally describe histopathological regions in IR image. Such a process is called prediction.. Patient pathology is finally recognized using Receiver Operating Characteristic (ROC) curves, based on the number of pixels (from a chosen class/classes) present in predicted images (Cancer class in the Patient pathology classification panel in Figure 1D). Model prediction power is evaluated using Area Under the Curve (AUC).

**Figure 1.**
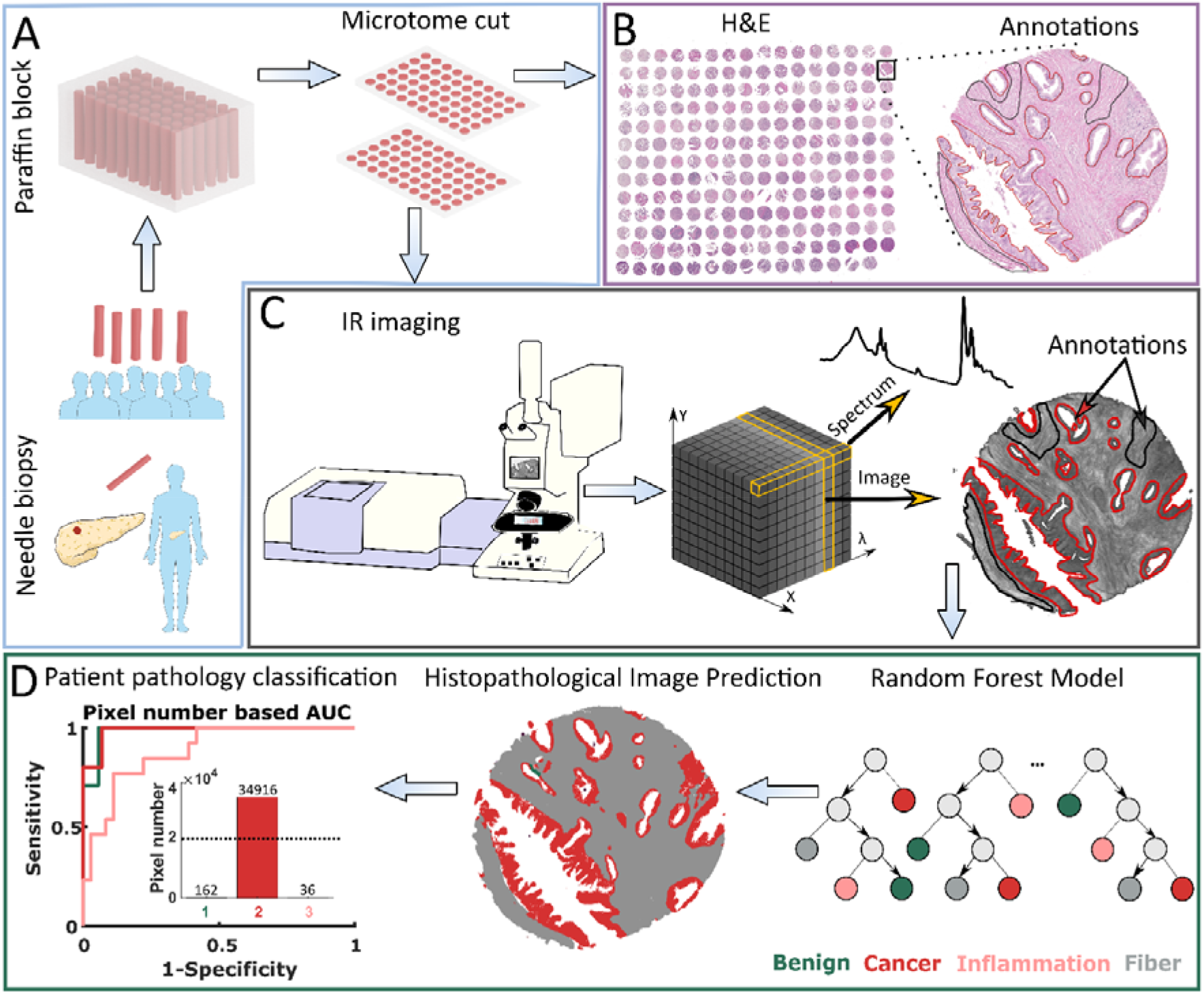
Patient pathology recognition pipeline: A) Tissue Micro Array preparation, B) Histopathological annotation on H&E stained biopsies images, C) Biopsies’ Infrared Imaging and obtained data structure, D) Model creation and patient pathology recognition.

### Comprehensive histopathological model

The FT-IR histopathological Random Forest model was based on 6 TMAs (more information in Supplementary Materials Table S1). Our Standard Definition based model can differentiate six classes of pancreatic tissue: Benign, Cancer, Necrosis, Inflammation, Fiber, and Blood. Differences between analyzed classes are to some extent reflected in their mean spectra (Figure 2B). At the first glance, there are visible differences in shape and relative intensity of Amide A and B (3550 – 3000 cm^-1^) protein bands and DNA/RNA bands (1300 – 1000 cm^-1^). Patients in TMAs were divided into model, validation, and test sets (Supplementary Materials Table 3) for model optimization and prediction power evaluation. In Figure 2A predicted TMAs (for Standard Definition) images with zooms on single cores and regions of interest are presented. Benign biopsies with green color can be easily differentiated from remaining cases in all TMAs. Class benign can be described as a combination of lobules and islands annotations, coming from normal pancreas tissue (for example single core from PA1002a TMA presented in Figure 2A), and normal adjacent to cancer tissue. The microenvironment of cancer is stroma-rich, which is visible among others in the single core from PA2072a TMA presented in Figure 2A. In both presented cancerous cores in the upper part of Figure 2A there can be also found necrotic regions associated with cancer changed biopsy tissue. Frequently, we observed lymphocyte clusters like in the single biopsy in PA961e TMA presented in Figure 2A, infiltrating into the cancerous tissue surrounded by stroma. Most chronic pancreatitis cases are visible in BBS14011 TMA. In the single biopsy predicted image, one may observe both: lymphocyte infiltration into the tissue and inflamed changed lobules. In the single core in BIC14011a TMA in Figure 2A, there is visible very well classified aggregation of erythrocytes in the affected stroma. The classification results on the pixel level (tissue level) were quantified using AUC metric in Figure 2C. All classes reached AUC values higher than 0.95, which is a very good result taking into account that AUC values equal to one corresponds to the situation where each pixel was correctly classified.

**Figure 2.**
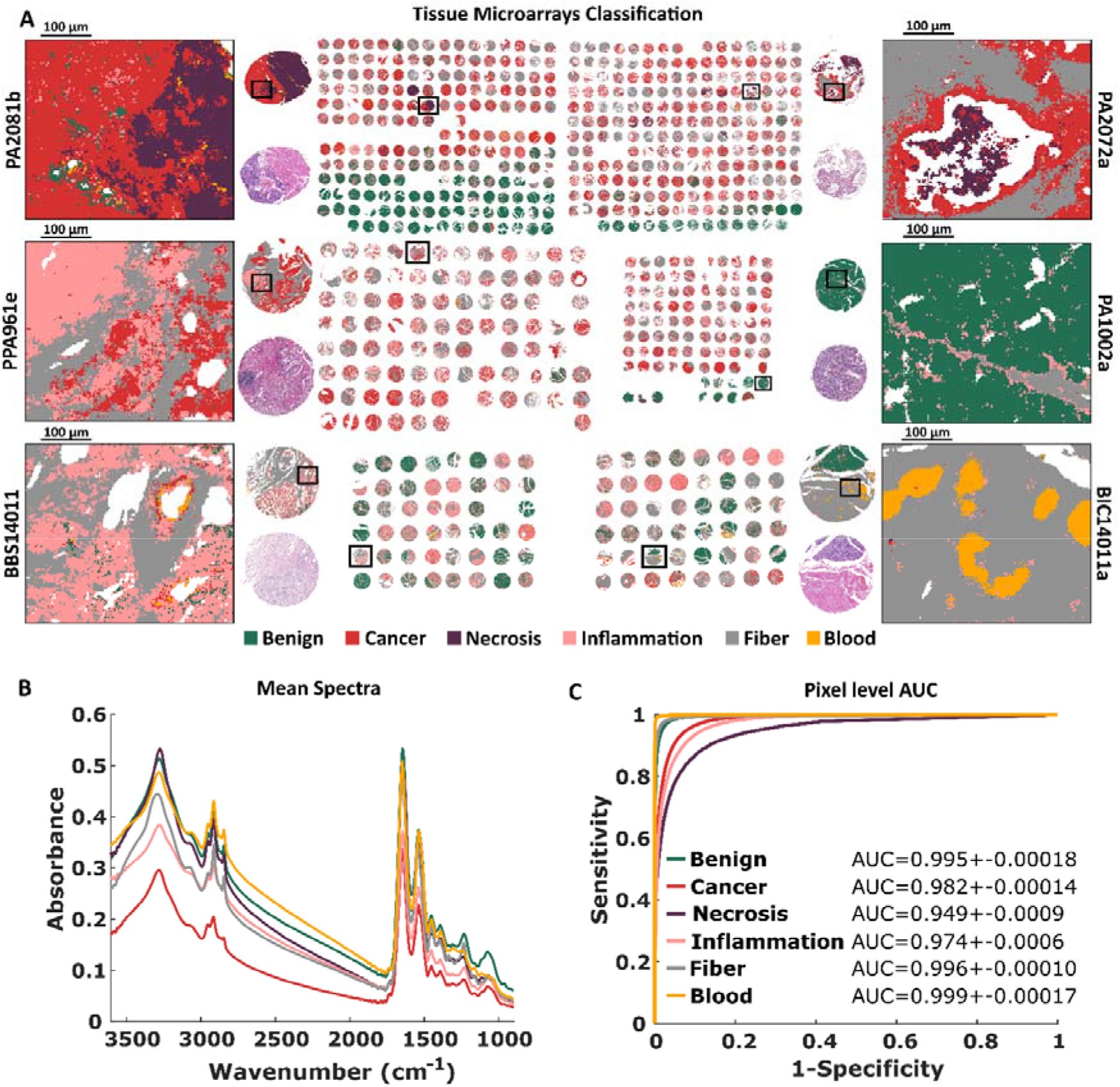
Standard Definition based histopathological recognition of six pancreatic tissue classes with Random Forest classification for: A) Tissue Micro Arrays with zooms on single cores with chosen regions, B) Tissue Micro Arrays mean spectra, and C) Pixel level AUC values of recognized tissue classes.

Finally, we used intraoperative biopsy (taken in clinical conditions) as the external sample for Standard Definition model assessment (Figure 3A). The most valuable result achieved for this sample is a very well classified transition from benign to cancerous state with inflammation infiltration. Zooms on interesting intraoperative biopsy regions are also presented (Figure 3B-D). In the predicted image in Figure 3 intraoperative margin can be easily marked.

**Figure 3.**
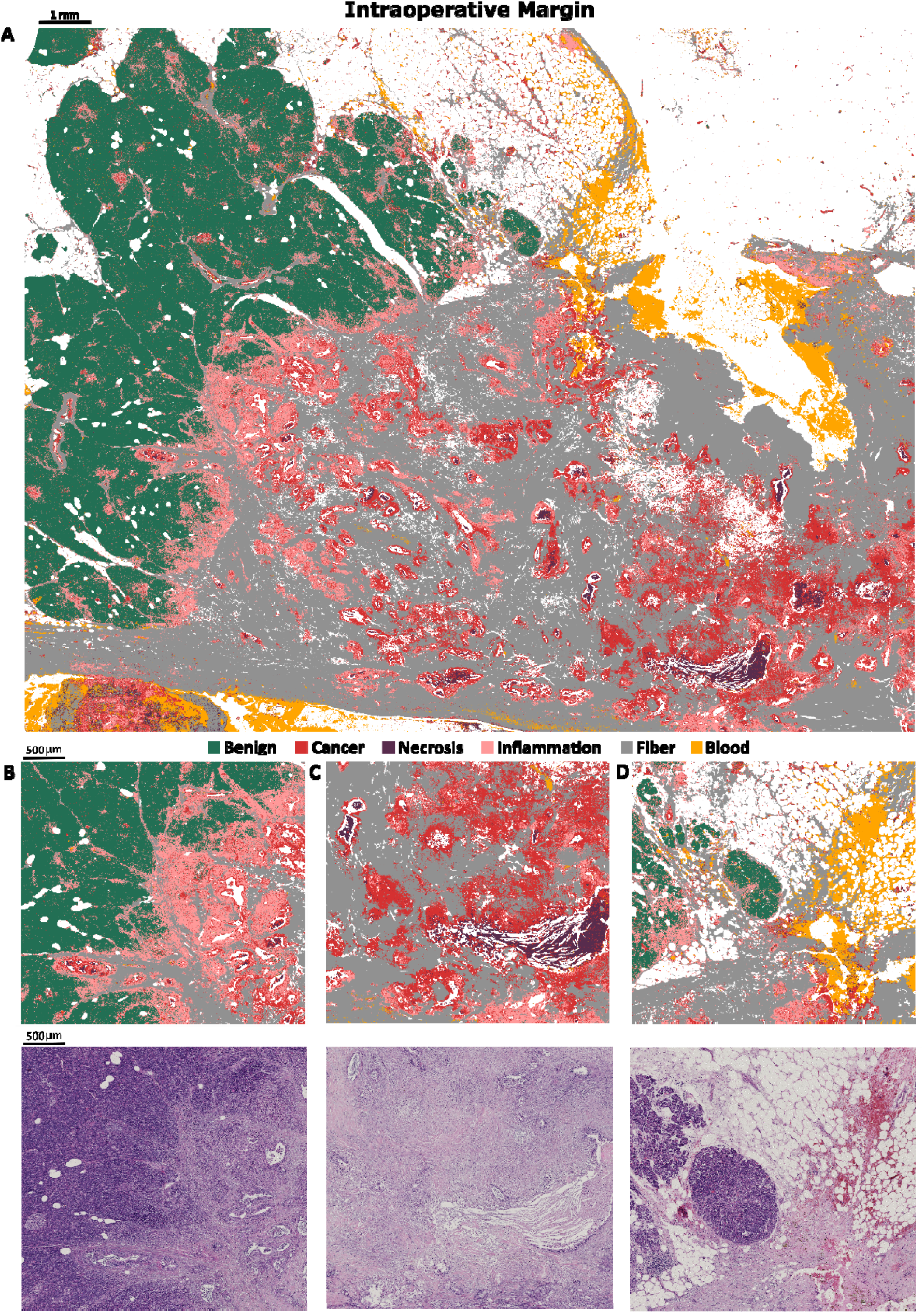
Predicted image of intraoperative biopsy A), with zooms on: B) margin between benign and cancerous tissue, with clearly visible inflammation between, C) big necrosis region surrounded by cancerous changed tissue, D) benign tissue with small inflammation and blood infiltration.

### High definition histopathological model

A similar model based on the same histopathological annotations and TMAs was built for High Definition data. The projected pixel size was 1.1 μm ensuring sufficient spatial sampling (for 0.5 NA objective) to retrieve the maximum of spatial information. In comparison to Standard Definition, this model unraveled small cells (i.e. lymphocytes) located between tissue. In general, predicted classes are not as consistent as in Standard Definition, there are cases with pixels that are highly mixed (Figure 4A). This is reflected in AUC values – there are three classes with results below 0.95 (Figure 4C). The mean spectra (and differences between them) for the HD data set (Figure 3B) are similar to those calculated for the SD data set in Figure 2B.

**Figure 4.**
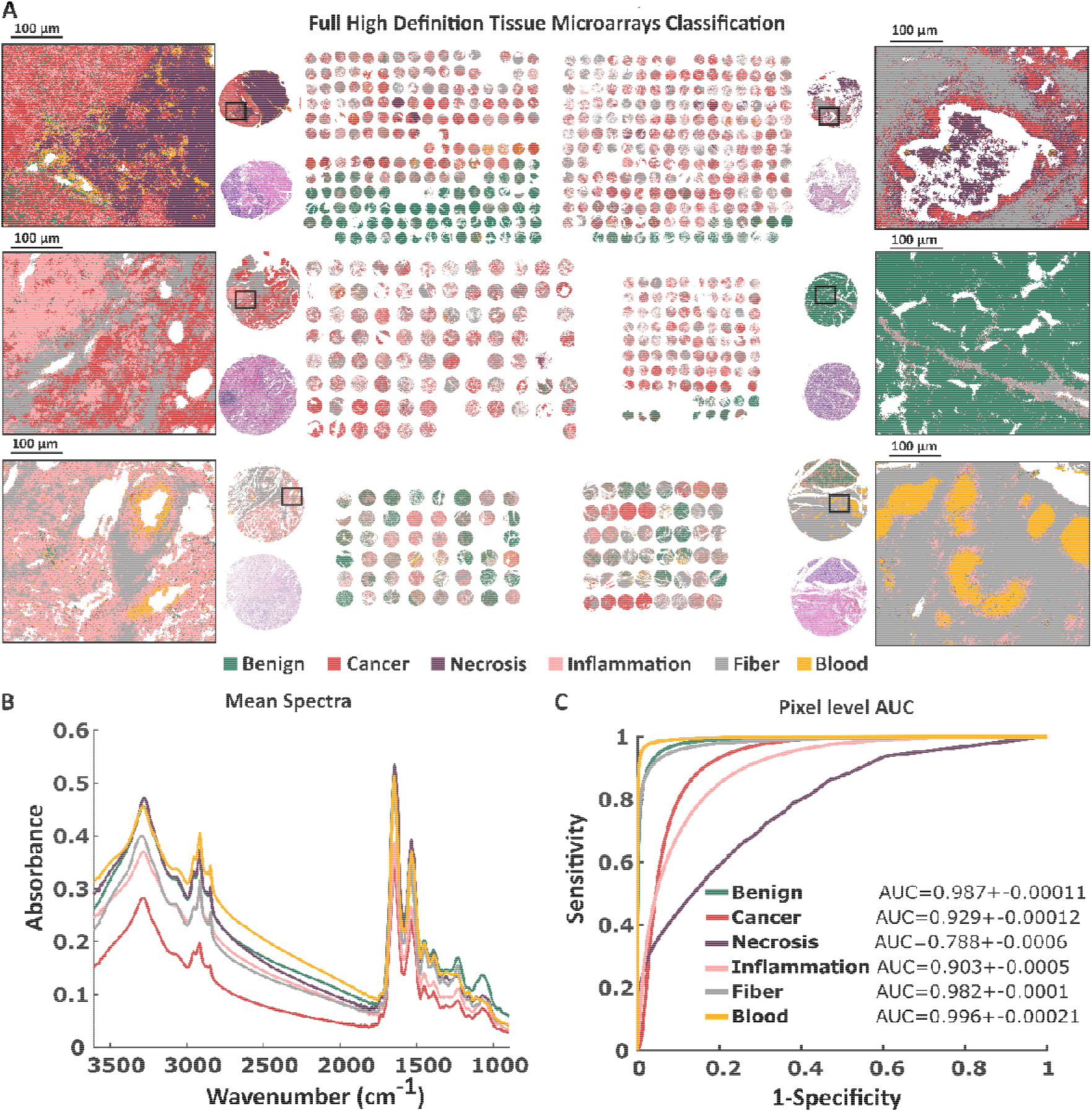
High Definition based histopathological recognition of 6 pancreatic tissue classes with Random Forest classification for A) Tissue Micro Arrays with zooms on single cores with chosen regions. B) Tissue Micro Arrays mean spectra, and C) Pixel level AUC values of recognized tissue classes.

We also calculated AUC metrics on the core and patient level for test set (49 patients) giving high AUC results in the range 0.99 (Figure 5) regardless of the definition.

**Figure 5.**
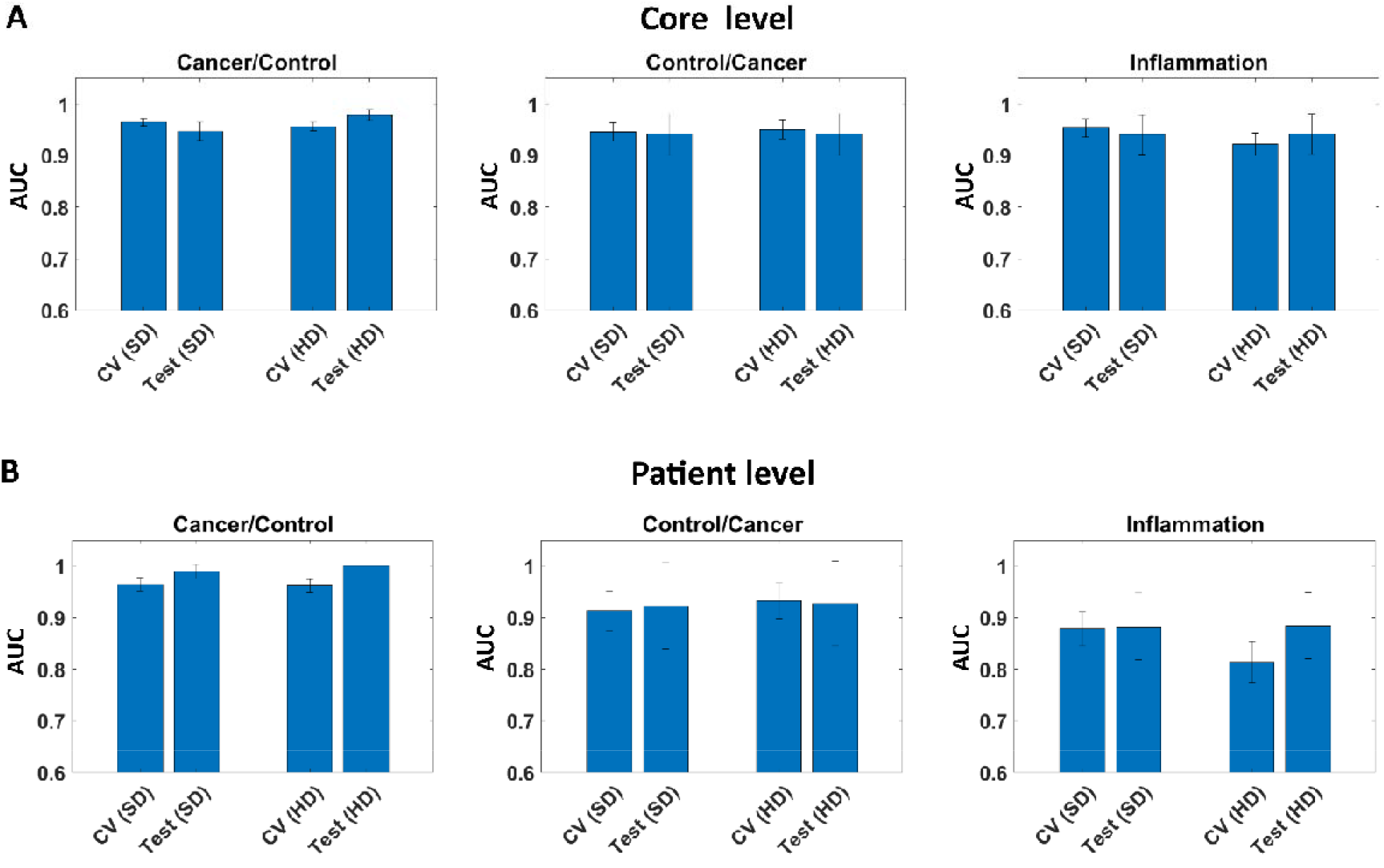
AUC for model (cross validation set - CV) and test set on: A) Core level, and B) Patient level for Standard (SD), and High Definitions (HD).

The Standard or High Definition images seem to provide a very similar contrast for this 6 class model. Similar results have been found for other pathologies [16] which means that for most cases High Definition is not required for a robust model. The increased noise level actually makes the models slightly less stable, while still having very high performance.

## Conclusions

The infrared imaging combined with the machine learning algorithm was applied to develop a model predicting comprehensive pancreatic histopathology. In the first step FT-IR Random Forest model was built based on a broad and reach in biochemical information spectra. This model has enabled recognition of six tissue classes: Benign, Cancer, Necrosis, Inflammation, Fiber, and Blood for both Standard and High Definition. In predictions based on High Definition data it was possible to differentiate inflammatory cells in more detail, however, predicted images were of better quality for Standard Definition. Pathology recognition that can be performed based on this model allows differentiation of cancerous, benign, and chronic pancreatitis cases with sensitivity higher that 95%. Moreover, in the event of a nonobvious case, an image of a predicted biopsy can be analyzed further to diagnose a patient finding specific regions of affected tissue, i.e. immune cells and erythrocyte infiltration, fibrosis, changes in a number of characteristic tissue structures. Model for Standard Definition achieved similar AUC values for cancerous patient detection to High Definition – that is positive in the context of research continuation using fast QCL system (that is equipped with objective giving projected pixel size down to 1.3 μm). This study opens the way for more subtle pathology recognition in pancreatic tissues.

## Methods

All calculations described in this study were done in MATLAB software.

### Biological material

Human pancreatic tissue was used in this study, with sample set consisting of six Tissue Microarrays (TMAs): PA2081b, PA2072a, PA1002a, PA961e, BBS14011, BIC14011a purchased from Biomax Inc. and two surgical resections. Since biopsy collection and personal data anonymization was done by Biomax Inc. no written consent from patients was requested. TMAs are assembled from tissue cores coming from needle biopsies with diameters between 1-1.5 mm (depending on the TMA). Resection material was specifically selected to present boarded between cancer and control tissue (surgical margin), therefore sample sizes are roughly 16x35 mm and 16x13 mm. Samples were placed in a paraffin block and cut using microtome for 6 μm thick slices, with one slice placed on IR transparent BaF_2_ salt plate for transmission measurement, and consecutive slice mounted on glass for Hematoxylin and Eosin (H&E) staining allowing histopathological annotations. Initially paraffin covered IR samples were deparaffinized with 24 hours hexan bath to avoid paraffin absorption around 1462 cm^-1^.

High patient statistic was achieved by the use of TMAs with 663 tissue cores in total. However, in most cases, more than one tissue core came from a single patient. Additionally, overlapping cases existed between some TMAs, thus, final patient cohort comprised of 250 cases (with two additional patients represented by surgical resection). Ethical approval was granted by Ethics committee at Jagiellonian University in Krakow (no. 1072.6120.304.2020).

### Fourier Transform Infrared Imaging

Bruker Vertex 70v spectrometer coupled with Hyperion 3000 microscope and 64x64 FPA MCT detector was used for FT-IR measurements. Spectra were measured within 3850-900 cm^-1^ range, with 8 cm^-1^ spectral resolution and zero filling factor of 1, giving 765 spectral points. Signal was co-averaged 4 times for a sample region and 64 times for background collection. Two types of objectives were used during measurements – 15x (SD) and 36x (HD), giving projected pixel sizes of 2.7 μm and 1.1 μm, respectively. All TMAs were measures with both resolutions, but surgical resections samples were only measured using 15x objective. Total number of sample spectra collected with FT-IR imaging in this study reached 672 407 632 (including both resolutions). Additional information on experimental time optimization is provided in Supplementary Materials.

The first step of preprocessing was data denoising using Minimum Noise Fraction (MNF) method with 20 bands used for reconstruction [17,18]. Due to partially spatial character of MNF’s noise estimation and RAM memory constrains, TMAs processing was done in a core-by-core manner. In case of resection material, denoising was done for region of 2x2 measurement tiles. Such approach allows to prevent information leakage between patients, which could cause over optimistic results in further classification. For the classification purpose, FT-IR spectra were transferred into so-called metrics. Metrics provide spectral information extracted from selected IR bands defined by experienced spectroscopist (list of spectral regions presented in Table S2 in Supplementary Materials). Metrics calculation for each spectral region includes local linear baseline removal followed by calculation of band integration, maximum value and center of gravity. Finally, those three characteristics are normalized to the corresponding value of Amide I. Total number of metrics extracted from single spectrum is 123.

### Random Forest Classification

Classification model creation requires multiple steps, starting from tissue types annotation and Region of Interests (ROIs) definition for chosen classes, which are required to extract data later used to feed the classifier. Annotations of Infrared images for all TMAs were done utilizing ground truth annotations provided by an experienced Histopathologist, based on visual inspection of H&E images. Such annotations were later transformed into ROI masks, allowing correct pixels (metrics/spectra) selection. Furthermore, to assure classification using only tissue originating pixels, a tissue mask was created after thresholding absorbance for 1650 cm^-1^ (most intensive band). Such a mask was used to remove background spectra from ROI masks and to select samples for final prediction images.

To prevent from Random Forest classifier being biased, all classes had the same number of samples (pixels) during a particular model training process. This number was determined by the number of samples available for the smallest class. Moreover, to ensure training data variability, equal number of samples from each TMA was taken, unless the number of available samples was insufficient. In such case, lacking samples were evenly topped up from other TMAs. In all cases, samples were always randomly selected.

FT-IR based six-class models (SD and HD) were built with 50 Trees, which was sufficient to prevent overtraining, but at the same time did not unnecessarily complicate the model. A crucial factor for stable model creation is its proper validation, assuring its stability and lack of overtraining. For FT-IR based models, patients were divided according to their annotations (benign, inflammation, cancer) in a random manner into model, validation and test sets. Subset of patients from each TMA was assigned to a test set, keeping in mind overlapping cases between TMAs. Number of patients in validation and test sets are presented in Supplementary Materials Table S3. A four-fold cross validation (using model data) was used during the model optimization, which is a long and iterative process. To assess models’ performance during the cross validation, confusion matrix along with Receiver Operating Characteristic (ROC) and Area Under the Curve (AUC) were calculated. Finally, when created model reached satisfactory parameters, a test set was used for its final evaluation with ROC and AUC defining final diagnostic ability. Described validation approach was applied to both, SD and HD datasets. Nonetheless, adopted validation method is strongly dependent from available patient cohort. Additional information on models validation is provided in Supplementary Materials.

## Data Availability

All data produced in the present study are available upon reasonable request to the authors

## Author Contributions

Study design: T.P.W., P.K., D.L.. Development of methodology: T.P.W., P.K., D.L.. Acquisition of spectroscopic data: T.P.W., P.K., D.L.. Data processing and analysis: P.K., D.L.. Writing the manuscript D.L., P.K., T.P.W..

## Acknowledgements

Authors wanted to acknowledge Slawka Urbaniak for histopathological annotations. This research was supported by „Pancreatic cancer comprehensive histopathology based on IR chemical imaging” project, which was carried out within the Homing programme (grant no. Homing/2016-2/20) of the Foundation for Polish Science co-financed by the European Union under the European Regional Development Fund. This research was performed using equipment purchased in the frame of the project co-funded by the Malopolska Regional Operational Program Measure 5.1 Krakow Metropolitan Area as an important hub of the European Research Area for 2007-2013, project no. MRPO.05.01.00-12-013/15.

## Data Availability

Data available on a reasonable request from the authors.

## References

[1] T.P. Wrobel, R. Bhargava, Infrared Spectroscopic Imaging Advances as an Analytical Technology for Biomedical Sciences, Anal. Chem. 90 (2018) 1444–1463. https://doi.org/10.1021/acs.analchem.7b05330.

[2] M.J. Baker, H.J. Byrne, J. Chalmers, P. Gardner, R. Goodacre, A. Henderson, S.G. Kazarian, F.L. Martin, J. Moger, N. Stone, J. Sulé-Suso, Clinical applications of infrared and Raman spectroscopy: State of play and future challenges, Analyst. 143 (2018) 1735–1757. https://doi.org/10.1039/c7an01871a.

[3] S. Mittal, T.P. Wrobel, M. Walsh, A. Kajdacsy-Balla, R. Bhargava, Breast cancer histopathology using infrared spectroscopic imaging: The impact of instrumental configurations, Clin. Spectrosc. 3 (2021) 100006. https://doi.org/10.1016/j.clispe.2021.100006.

[4] S. Tiwari, A. Kajdacsy-Balla, J. Whiteley, G. Cheng, S.M. Hewitt, R. Bhargava, INFORM: INFrared-based ORganizational Measurements of tumor and its microenvironment to predict patient survival, Sci. Adv. 7 (2021). https://doi.org/10.1126/sciadv.abb8292.

[5] F. Großerueschkamp, A. Kallenbach-Thieltges, T. Behrens, T. Brüning, M. Altmayer, G. Stamatis, D. Theegarten, K. Gerwert, Marker-free automated histopathological annotation of lung tumour subtypes by FTIR imaging, Analyst. 140 (2015) 2114–2120. https://doi.org/10.1039/c4an01978d.

[6] J.R. Hands, P. Abel, K. Ashton, T. Dawson, C. Davis, R.W. Lea, A.J.S. McIntosh, M.J. Baker, Investigating the rapid diagnosis of gliomas from serum samples using infrared spectroscopy and cytokine and angiogenesis factors, Anal. Bioanal. Chem. 405 (2013) 7347–7355. https://doi.org/10.1007/s00216-013-7163-z.

[7] J.T. Kwak, A. Kajdacsy-Balla, V. Macias, M. Walsh, S. Sinha, R. Bhargava, Improving prediction of prostate cancer recurrence using chemical imaging, Sci. Rep. 5 (2015) 1–10. https://doi.org/10.1038/srep08758.

[8] D. Liberda, M. Hermes, P. Koziol, N. Stone, T.P. Wrobel, Translation of an esophagus histopathological FT-IR imaging model to a fast quantum cascade laser modality, J. Biophotonics. 13 (2020) 1–9. https://doi.org/10.1002/jbio.202000122.

[9] M. Schnell, S. Mittal, K. Falahkheirkhah, A. Mittal, K. Yeh, S. Kenkel, A. Kajdacsy-Balla, P.S. Carney, R. Bhargava, All-digital histopathology by infrared-optical hybrid microscopy, Proc. Natl. Acad. Sci. U. S. A. 117 (2020) 3388–3396. https://doi.org/10.1073/pnas.1912400117.

[10] S. Mittal, K. Yeh, L. Suzanne Leslie, S. Kenkel, A. Kajdacsy-Balla, R. Bhargava, Simultaneous cancer and tumor microenvironment subtyping using confocal infrared microscopy for all-digital molecular histopathology, Proc. Natl. Acad. Sci. U. S. A. 115 (2018) E5651–E5660. https://doi.org/10.1073/pnas.1719551115.

[11] C. Kuepper, A. Kallenbach-Thieltges, H. Juette, A. Tannapfel, F. Großerueschkamp, K. Gerwert, Quantum Cascade Laser-Based Infrared Microscopy for Label-Free and Automated Cancer Classification in Tissue Sections, Sci. Rep. 8 (2018) 1–10. https://doi.org/10.1038/s41598-018-26098-w.

[12] N. Goertzen, R. Pappesch, J. Fassunke, T. Brüning, Y.D. Ko, J. Schmidt, F. Großerueschkamp, R. Buettner, K. Gerwert, Quantum Cascade Laser-Based Infrared Imaging as a Label-Free and Automated Approach to Determine Mutations in Lung Adenocarcinoma, Am. J. Pathol. 191 (2021) 1269–1280. https://doi.org/10.1016/j.ajpath.2021.04.013.

[13] K. Szymoński, E. Lipiec, K. Sofińska, K. Skirlińska-Nosek, K. Milian-Ciesielska, J. Szpor, M. Czaja, S. Seweryn, N. Wilkosz, G. Birarda, F. Piccirilli, L. Vaccari, M. Szymoński, Spectroscopic screening of pancreatic cancer, Clin. Spectrosc. 3 (2021) 100016. https://doi.org/10.1016/j.clispe.2021.100016.

[14] V. Notarstefano, S. Sabbatini, C. Conti, M. Pisani, P. Astolfi, C. Pro, C. Rubini, L. Vaccari, E. Giorgini, Investigation of human pancreatic cancer tissues by Fourier Transform Infrared Hyperspectral Imaging, J. Biophotonics. 13 (2020) 1–10. https://doi.org/10.1002/jbio.201960071.

[15] D. Liberda, K. Kosowska, P. Koziol, T.P. Wrobel, Spatial sampling effect on data structure and Random Forest classification of tissue types in High Definition and Standard Definition FT-IR imaging, Chemom. Intell. Lab. Syst. 217 (2021) 104407. https://doi.org/10.1016/j.chemolab.2021.104407.

[16] S. Mittal, T.P. Wrobel, M. Walsh, A. Kajdacsy-Balla, R. Bhargava, Breast cancer histopathology using infrared spectroscopic imaging: The impact of instrumental configurations, Clin. Spectrosc. 3 (2021) 100006. https://doi.org/10.1016/j.clispe.2021.100006.

[17] M.K. Raczkowska, P. Koziol, S. Urbaniak, C. Paluszkiewicz, W.M. Kwiatek, T.P. Wrobel, Influence of denoising on classification results in the context of hyperspectral data□: High Definition FT-IR imaging, Anal. Chim. Acta. 1085 (2019) 39–47.

[18] P. Koziol, M.K. Raczkowska, J. Skibinska, S. Urbaniak-Wasik, C. Paluszkiewicz, W. Kwiatek, T.P. Wrobel, Comparison of spectral and spatial denoising techniques in the context of High Definition FT-IR imaging hyperspectral data, Sci. Rep. 8 (2018) 1–11. https://doi.org/10.1038/s41598-018-32713-7.

